# Cerebrospinal fluid purinomics as a biomarker approach to predict outcome after severe traumatic brain injury

**DOI:** 10.1101/2021.10.20.21265297

**Authors:** Nathan Ryzewski Strogulski, Marco Antonio Stefani, Ana Elisa Böhmer, Gisele Hansel, Marcelo S. Rodolphi, Afonso Kopczynski, Vitória Girelli de Oliveira, Eduarda Tanus Stefani, Juliana Vinadé Portela, André P. Schmidt, Jean Pierre Oses, Douglas H. Smith, Luis V. Portela

## Abstract

Severe traumatic brain injury (TBI) is associated with high rates of mortality and long-term disability linked to neurochemical abnormalities. Although purine-derivatives play important roles in TBI pathogenesis in preclinical models, little is known about potential changes in purine levels and their implications in human TBI. We assessed cerebrospinal fluid (CSF) levels of purines in severe TBI patients as potential biomarkers that predict mortality and long-term dysfunction. This was a cross-sectional study performed in 17 severe TBI patients (Glasgow Coma Scale < 8) and 51 controls. Two to four hours after admission to ICU, patients were submitted to ventricular drainage, and CSF collection for quantification of adenine and guanine purine-derivatives by HPLC. TBI patients survival was followed up to 3 days from admission. A neurofunctional assessment was performed through modified Rankin Scale (mRS) two years after ICU admission. Purine levels were compared between control and TBI patients, and between surviving and non-surviving patients. Relative to controls, TBI patients presented increased CSF levels of GDP, guanosine, adenosine, inosine, hypoxanthine, and xanthine. Further, GTP, GDP, IMP, and xanthine levels were different between surviving and non-surviving patients. Among the purines, guanosine was associated with improved mRS (p=0.042; r= −0.506). Remarkably, GTP displayed predictive value (AUC=0.841, p=0.024) for discriminating survival vs. non-survival patients up to three days from admission. These results support TBI-specific purine signatures, suggesting GTP as a promising biomarker of mortality, and guanosine as an indicator of long-term functional disability.

**Highlights:**

- CSF levels of guanosine, GDP, adenosine, inosine, hypoxanthine, and xanthine are increased in severe TBI patients.
- GTP, GDP, IMP and xanthine levels were different between surviving and non-surviving patients.
- Guanosine was associated to improved neurological outcomes two years after TBI.
- CSF GTP levels at admission predicted patient death.

## Introduction

Traumatic brain injury (TBI) is considered a major health concern with a high incidence of disability worldwide, and it is the leading cause of mortality among young adults from low- and middle-income countries (Nguyen et al., 2016). In severe TBI, immediate mechanical damage to brain tissue can be life-threatening (Braun CT, 2015) and can trigger a series of secondary neurochemical cascades that further damage the brain. Some of the most well characterized of these processes include excitotoxicity, formation of proteotoxic aggregates, neuroimmune dysfunction, and changes in brain metabolism (Doran et al., 2019; Henry et al., 2020; Johnson et al., 2013, 2012; Lyons et al., 2018; Stefani et al., 2017). However, while adenine- and guanine-based purines have been shown to play important roles in the pathogenesis of other brain disorders, little is known about their contribution to the post-TBI sequelae.

Beyond their central role in formation of nucleic acids and bioenergetics, purines have a wide range of neuromodulatory effects that are disrupted in various neurological disorders. Indeed, purinergic receptors have been explored as therapeutic targets in animal models of epilepsy, amyotrophic lateral sclerosis, Alzheimer’s, and Parkinson’s disease (Burnstock, 2020, 2017). An important feature of normal purine-mediated brain responses is the strict control of their individual concentrations, performed by intra and extracellular enzymes (Robson et al., 2006). In particular, the Ecto-NTPDases are a family of enzymes responsible for the extracellular degradation of purines, thereby controlling the binding probability to specific purinergic receptors and the magnitude of receptor activation (Robson et al., 2006). This includes the stepwise extracellular degradation of ATP and GTP, resulting in the formation of ADP and AMP, or GDP and GMP, respectively. Further catabolism leads to correspondent nucleosides adenosine and guanosine, which consequently generate IMP, inosine, hypoxanthine, xanthine, and uric acid (Cunha, 2016; Pelligrino et al., 2010; Zeng et al., 2018). Purine-derived metabolites also cross plasma membranes through specific transporters, influencing their intra- and extracellular levels (Beal et al., 2004). Remarkably, from the extracellular triphosphate forms to their downstream metabolites, each purine derivative may exert characteristic physiological roles in the brain mediated by purinergic receptors and GTPase-activating proteins, neuroinflammatory effectors, and glutamatergic neurotransmission. Hence, changes in the purine levels in the brain after TBI could play an important role in outcomes from TBI.

While clinical studies of purines in TBI have been limited, Headrick and colleagues demonstrated that acute increases in extracellular adenosine levels were associated with impaired neuroenergetic and neurological functions in a preclinical rat TBI model (Headrick et al., 1994). In addition, inhibition of adenosinergic signaling with caffeine was shown to decrease the mortality rate of rats submitted to severe TBI (Lusardi et al., 2012). Furthermore, intraperitoneal administration of guanosine after experimental TBI attenuated cognitive and mitochondrial dysfunction (Dobrachinski et al., 2019; Gerbatin et al., 2017). Finally, *in vivo*, extracellular levels of adenosine, inosine, and hypoxanthine were found to increase soon after TBI, which was mechanistically linked to energy failure mirrored by the rapid ATP catabolism to adenosine (Bell et al., 1998). Whereas these few experimental studies reveal potential mechanisms of purine metabolism in TBI, the potential clinical relevance of extracellular purine levels as prognostic biomarkers had received little attention.

In this study, we assessed cerebrospinal fluid (CSF) levels of purines in severe TBI patients to identify specific signatures associated with mortality and long-term neurological disfunction.

## Subjects and Methods

### Study population and clinical management

This is a single center cross-sectional study, carried out in the Emergency Unit of the Cristo Redentor Hospital (Porto Alegre, RS, Brazil). This study included, a total of 17 severe TBI patients already described previously, presenting on hospital admission both Glasgow Coma Scale (GCS) ≤ 8 and an abnormal brain CT scan (Böhmer et al., 2011; Stefani et al., 2017). Patients were predominantly male (mean [SD] age, 29 [13] years; M/F, 9:1). Inclusion criteria was an isolated severe TBI, with no previous history of comorbidities that could influence the biomarkers’ concentrations and clinical outcomes. Exclusion criteria, and clinical management were previously reported (Böhmer et al., 2011; Stefani et al., 2017). CSF was collected between 2 and 4 h after hospitalization through an intraventricular catheter.

Additionally, 51 healthy subjects (ASA I status) scheduled for elective urological, gynecological, general, or vascular procedures were selected as age and sex-matched controls, also previously described (Böhmer et al., 2011). Experienced anesthesiologists collected the CSF after successful subarachnoid puncture and before the intrathecal injection of anesthetics or analgesics. The first 0.5 mL of CSF aspirated was discarded, and the subsequent 0.5 mL sample collected and inspected visually for blood contamination.

Immediately after CSF collection, the samples from controls and patients were centrifuged at 10,000 x g for 5 min to obtain a supernatant free of cells and cellular debris. The supernatant was then transferred to sealed plastic tubes, and stored at −70°C within 30 min of collection. Between 38 to 48 hours from collection, CSF samples were thawed over ice for purines assessment.

Informed consent for participating in this study was obtained from patients’ family members and directly from healthy individuals, according to the Declaration of Helsinki. Concomitantly, family members were questioned about patient’s lifestyle and pre-existent diseases. The local institutional Ethics Committee approved this protocol (project number 0038.0.164.165-05).

### Outcome measures

Patient outcome was defined accordingly: deterioration to brain death (non-survival n = 6) or survival (survival, n = 11), within 3 days after hospital admission. Deterioration to brain death occurred only up to 3 days after admission to the ICU. The ICP, hemodynamic, and metabolic variables including mean arterial blood pressure and cerebral perfusion pressure were assessed daily and reported previously (Böhmer et al., 2011). Two years after discharge, telephone calls were placed to confirm survival, and investigate the level of long-term functional disability. TBI patients were then assessed for the modified Rankin Scale (mRS) and scored by an experienced neurologist. The mRS ranks disability posterior to stroke and cerebral injuries, ranging from 6 (dead) to 0 (fully independent), and is considered a reliable endpoint for clinical neurological studies.

### CSF Purinomics

We carried out measures in 10µL CSF aliquots at a Shimadzu Class-VP chromatography system in which the separation of adenine and guanine based purines was achieved with a Supelco C18 250 column, as previously described by our group (Domanski et al., 2006; Oses et al., 2007; Schmidt et al., 2015). Absorbance was read at 254 nm, and quantification of all purines from each subject was obtained in a single run.

The following purines and metabolites were determined: adenosine triphosphate (ATP), adenosine diphosphate (ADP), adenosine monophosphate (AMP), adenosine (ADO), guanosine triphosphate (GTP), guanosine diphosphate (GDP), guanosine monophosphate (GMP), guanosine (GUO), inosine monophosphate (IMP), inosine (INO), hypoxanthine (HXN), xanthine (XAN), and uric acid (UA). The HPLC analysis was conducted by blinded researchers to the groups. The acquired data was stored in a data bank for future comparisons.

### Statistical analysis

Prior to perform statistical analysis, purines level of controls and patients were screened for mathematical outliers. Subjects that presented purines value above 1.5-fold of interquartile range from median, or 3-fold of standard deviation from mean were excluded from all analysis (Mowbray et al., 2019). Data obtained were then submitted to Kolmogorov-Smirnov testing for normality. Data was analyzed using RStudio (1.2.5001), and associated packages further indicated. Data is presented as mean ± SD, or median ± IQR, and the graphical representation was made using GraphPad Prism 8. Comparison between groups was performed by Student’s independent t test, or Mann-Whitney test. Reciever-Operator Curves (ROC) were created to explore the ability of biomarkers to predict survival, indicated by the Area Under the Curve (AUC). The cutoff value was calculated by the Youden method, using the R package “*OptimalCutpoints”* (López-Ratón et al., 2014). Multivariate correlations between purines, and clinical data was assessed through Spearman correlation test, with corrections by Bonferroni, using the R package “*Psych”* (Revelle, 2018) for statistical testing, and “*Corrplot”* (Wei and Simko, 2017) for graphical representation. The multivariate correlation of purine derivatives was utilized to create a model, which is based on the classical cascade of purine degradation obtained from KEGG database (Kanehisa, 2019; Kanehisa and Goto, 2000). The statistical significance was considered when *p* ≤ 0.05

## Results

### Severe TBI triggers specific changes in CSF purine metabolites

Among guanine-derived purines, GDP and guanosine were significantly increased in TBI patients, compared to controls (Figure 1A and B, and Table 1), while no difference was observed in GTP and GMP levels (Supplemental Figure 1A and B). In non-surviving TBI patients, there was increased CSF concentrations of GTP, and GDP compared with surviving patients (Figure 1C and D, respectively). Concentrations of GMP and guanosine did not differ between surviving and non-surviving patients (Table 1, supplemental figure 1C and D).

**Table 1:** Descriptive data of studied population, comparing TBI patients and matched controls, and surviving and non-surviving TBI patients. Descriptive statistics are presented as mean ± SD, or median (Q1 – Q3), and p-values are reported for independent t-test, or Mann-Whitney U test, as p-value ([degrees of freedom] t-value), or p-value (Mann-Whitney U) respectively. * indicates p-values <0.05, ** indicates p-values <0.01 and *** indicates p-values < 0.001. Abbreviations: sTBI, Severe Traumatic Brain Injury; GCS, Glasgow Coma Score; ICP, Intracranial Pressure.

| Variables | Groups |  |  | Groups |  |  |
| --- | --- | --- | --- | --- | --- | --- |
|  | Controls<br>n=51 | sTBI Patients<br>n = 17 | p-value<br>([df]t/ U) | Surviving<br>n=11 | Non-surviving<br>n=6 | p-value<br>([df]t/ U) |
| **Age** | - | 29.89 ± 11.7 | - | 31.77 ± 12.9 | 24.6 ± 8.7 | 0.187<br>([15] -1.41) |
| **GCS at admission** | - | 6<br>(4 – 7) | - | 6<br>(6 – 7) | 4<br>(4 – 7) | 0.3637 (25.5) |
| **ICP at admission** | - | 12.89 ± 11.92 | - | 11.85 ± 11.05 | 14.2 ± 16.0 | 0.8265<br>([15] 0.203) |
| **GTP** | 2.140<br>(1.60 – 2.34) | 1.580<br>(1.24 – 2.65) | 0.2241<br>(347) | 1.505 ± 0.452 | 3.178 ± 1.74 | **0.0077<br>([15] 3.075) |
| **GDP** | 0.050<br>(0.000 – 0.120) | 0.470<br>(0.255 – 0.825) | ***<0.0001<br>(136.5) | 0.397 ± 0.28 | 0.819 ± 0.654 | *0.0431<br>([15] 2.209) |
| **GMP** | 0.000<br>(0.000 – 0.000) | 0.000<br>(0.000 – 0.000) | 0.9408<br>(454) | 0.000<br>(0.000 – 0.000) | 0.000<br>(0.000 - 1.170) | 0.353<br>(27.5) |
| **Guanosine** | 0.000<br>(0.000 – 0.0200) | 0.570<br>(0.430 – 1.965) | ***<0.0001<br>(0) | 0.740<br>(0.540 – 1.560) | 0.485<br>(0.338 – 3.048) | 0.524<br>(26) |
| **ATP** | 0.000<br>(0.000 – 0.000) | 0.000<br>(0.000 – 0.040) | 0.1073<br>(353) | 0.000<br>(0.000 – 0.040) | 0.000<br>(0.000 – 0.088) | >0.999<br>(32.50) |
| **ADP** | 0.170<br>(0.000 – 0.240) | 1.130<br>(0.865 – 1.330) | ***<0.0001<br>(50.50) | 1.138 ± 0.362 | 1.008 ± 0.720 | 0.6232<br>([15] 0.5017) |
| **AMP** | 1.710<br>(0.000 – 2.680) | 0.830<br>(0.520 – 2.370) | 0.6883<br>(403) | 0.830<br>(0.570 – 2.220) | 0.880<br>(0.298 – 2.620) | 0.7885<br>(30) |
| **Adenosine** | 0.050<br>(0.00 – 0.290) | 0.990<br>(0.560 – 2.040) | ***<0.0001<br>(37) | 1.210<br>(0.560 – 1.880) | 0.655<br>(0.545 – 5.028) | 0.9812<br>(35.20) |
| **IMP** | 1.440<br>(0.970 – 2.050) | 0.000<br>(0.000 – 1.500) | **<0.01 (252) | 0.000<br>(0.000 – 0.790) | 1.915<br>(0.000 – 3.053) | *0.0438<br>(15.5) |
| **Inosine** | 1.930 ± 0.855 | 2.711 ± 1.933 | *0.0241<br>([66] 2.308) | 3.124 ± 2.200 | 1.953 ± 1.100 | 0.2449<br>([15] 1.210) |
| **Xanthine** | 4.250<br>(3.290 – 8.340) | 8.330<br>(6.430 – 11.19) | **0.0037<br>(232) | 7.943 ± 2.076 | 16.96 ± 12.97 | *0.0352<br>([15] 2.315) |
| **Hypoxanthine** | 2.890<br>(2.210 – 3.660) | 5.510<br>(3.740 – 8.335) | ***<0.0001<br>(116) | 5.260<br>(3.340 – 7.660) | 6.805<br>(3.705 – 33.15) | 0.4043<br>(24) |
| **Uric Acid** | 24.35<br>(18.03 – 33.76) | 22.41<br>(15.13 – 31.08) | 0.5091<br>(386) | 25.95<br>(15.58 – 27.95) | 20.01<br>(12.50 – 129.7) | 0.9612<br>(32) |

**Figure 1.**
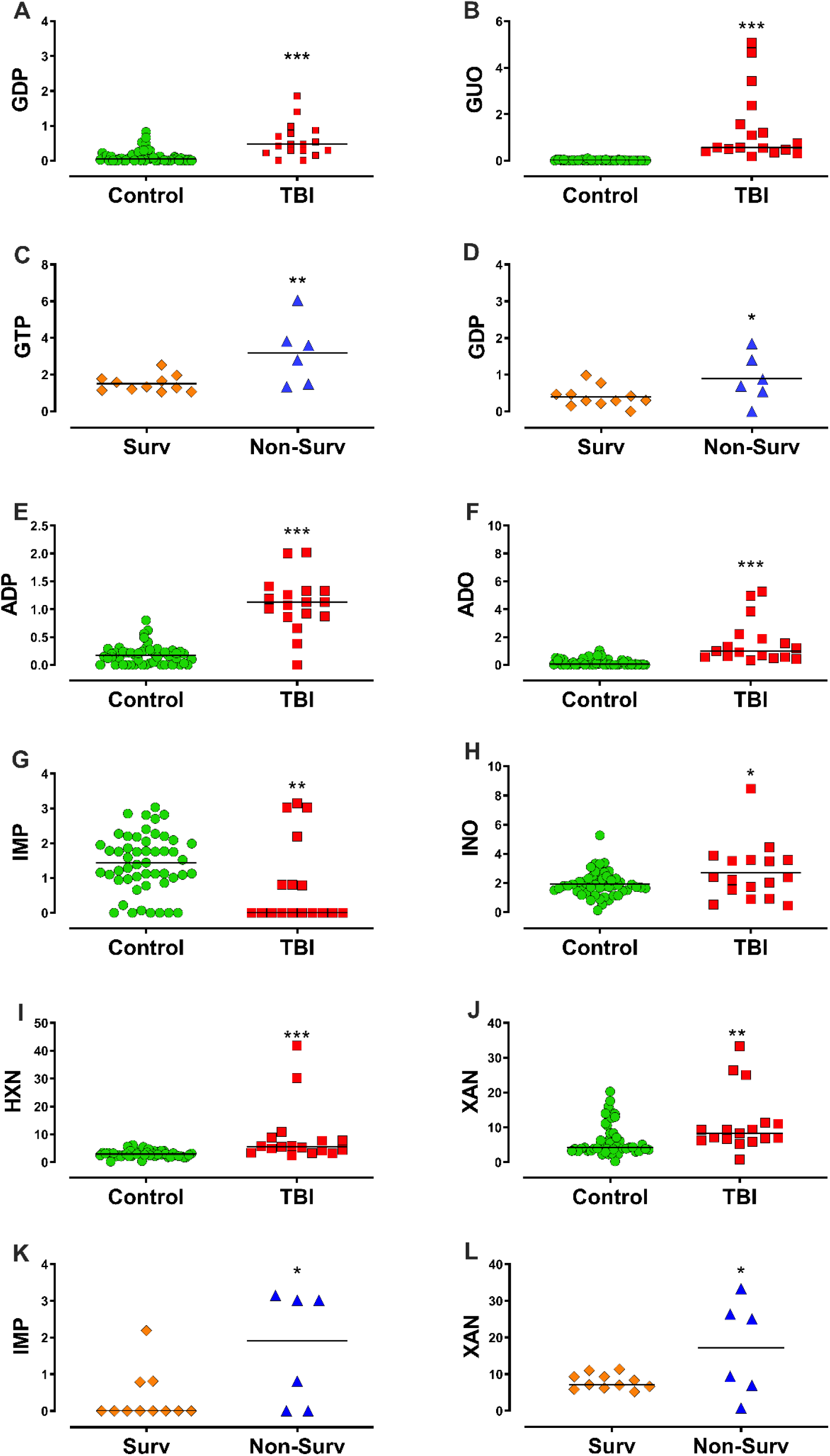
Cerebrospinal fluid levels of purines in control and TBI (surviving and non-surviving). This setting displays the composite of purines with statistically significant difference between groups. **(A)** Guanine derivatives GDP, **(B)** and guanosine (GUO) presented increased levels in TBI patients compared to controls, **(C)** and GTP **(D)** and GDP were increased in non-surviving (Non-Surv), relative to surviving patients (Surv). **(E)** Adenine derivatives ADP and **(F)** adenosine (ADO) were increased in TBI patients compared to controls. **(G)** CSF IMP levels were reduced in TBI patients compared to control, **(H)** whilst inosine (INO), **(I)** hypoxanthine (HXN) and **(J)** xanthine (XAN) were increased. Non-surviving patients presented increased IMP **(K)** and xanthine **(L)** CSF levels compared to surviving patients. Horizontal lines indicate median or mean. Statistical significance was assessed through Mann-Whitney or Student’s independent T test, with *, **, *** indicating *p* ≤ 0.05, *p* ≤ 0.01, *p* ≤ 0.001, respectively (Controls n= 51, TBI n = 17; Surv n =11, Non-Surv n=6).

Adenine-based purines, ADP and adenosine, were increased in TBI patients, relative to controls (Figure 1E and F, respectively, and Table 1), whilst no difference was observed in ATP and AMP levels (Supplemental figure 1E and F). Further, ATP, ADP, AMP, and adenosine were not different between surviving and non-surviving patients (Table 1, supplemental figure 1G to J).

The downstream guanine and adenine purine metabolism converge to the formation of IMP, inosine, hypoxanthine, xanthine, and uric acid. While CSF levels of IMP were reduced in TBI patients, inosine, hypoxanthine, and xanthine were increased (Figure 1G to J, respectively, also Table 1). No significant difference was observed in uric acid CSF levels (supplemental figure 1K). Among TBI patients, IMP and xanthine levels were increased in non-surviving compared with surviving patients (Figure 1K and L). Similar levels of inosine, hypoxanthine and uric acid were found in both surviving and non-surviving patients (supplemental figure 1L, M and N, respectively).

### CSF purinomics yield prognostic signatures for neurological outcomes

Considering that four different purine metabolites presented statistical difference between surviving and non-surviving TBI patients, we investigated the accuracy of these metabolites as prognostic biomarkers for mortality due to severe TBI.

The AUROC indicates how well GTP, GDP, IMP, and xanthine can discriminate patients that will survive from patients that will die up to 3 days from the admission to ICU (Figure 2A to D). With a cut-off value of > 2.780, GTP provided 66.67% of sensitivity, and 100% of specificity (AUROC: 0.841, *p* = 0.0237, 95% CI, CL: 95.45% to 100%). GDP, IMP, and xanthine did not present statistically significant predictive values (*p* > 0.05). However, the AUROC values for IMP barely reached significance (p = 0.06).

**Figure 2.**
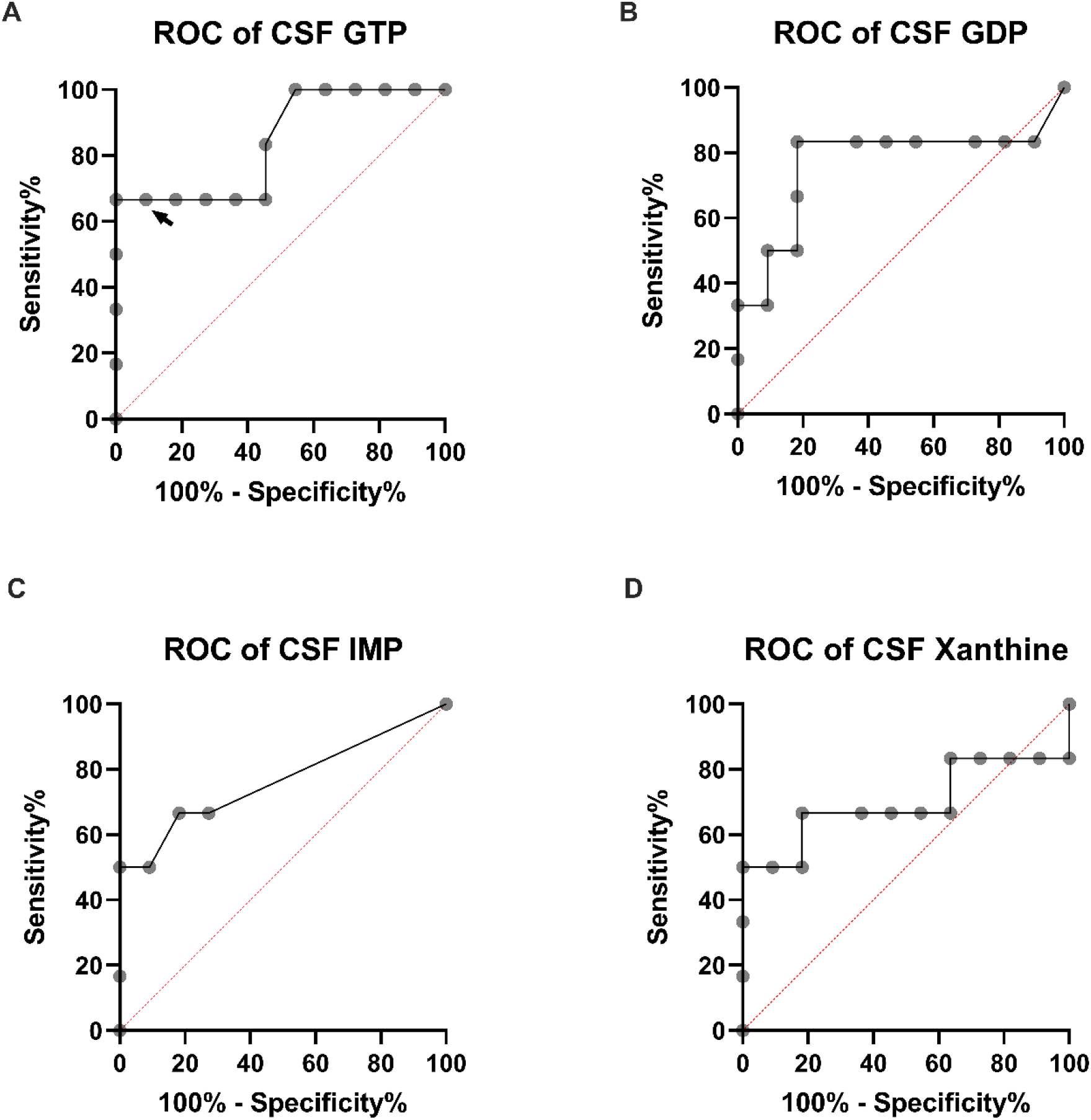
Predictive accuracy of CSF purines relative to death up to 3 days from ICU admission. Area under the curve of the receiver-operator characteristic curve (AUROC) for GTP **(A)**, GDP **(B)**, IMP **(C)** and Xanthine **(D). (A)** GTP presented at a cut-off value of >2.780 (indicated by arrowhead) a sensitivity of 66.67%, and 100% of specificity (AUROC: 0.841, *p* = 0.0237, 95% CI, CL: 62.26% to 99.53%, Youden Index for >2.780 = 0.667). CSF GDP **(B)**, IMP **(C)** and xanthine **(D)** levels presented AUROC with lower predictive value (*p* > 0.05) (Controls n= 51, TBI n = 17; Survival n =11, Non-Survival n=6).

Also, we performed bivariate correlations between CSF purine levels, and clinical neurological outcomes. We found that the modified Rankin Scale (mRS) did not correlate with TBI severity at admission (GCS) (*r*: −0.32, *p* = 0.207), whereas guanosine correlated with mRS (*r*: −0.50, *p* = 0.042) (Figure 3 A and B, respectively). Clinical parameters measured at ICU such as the medium arterial pressure, and cerebral perfusion pressure were not statistically correlated to mRS (Data not shown).

**Figure 3.**
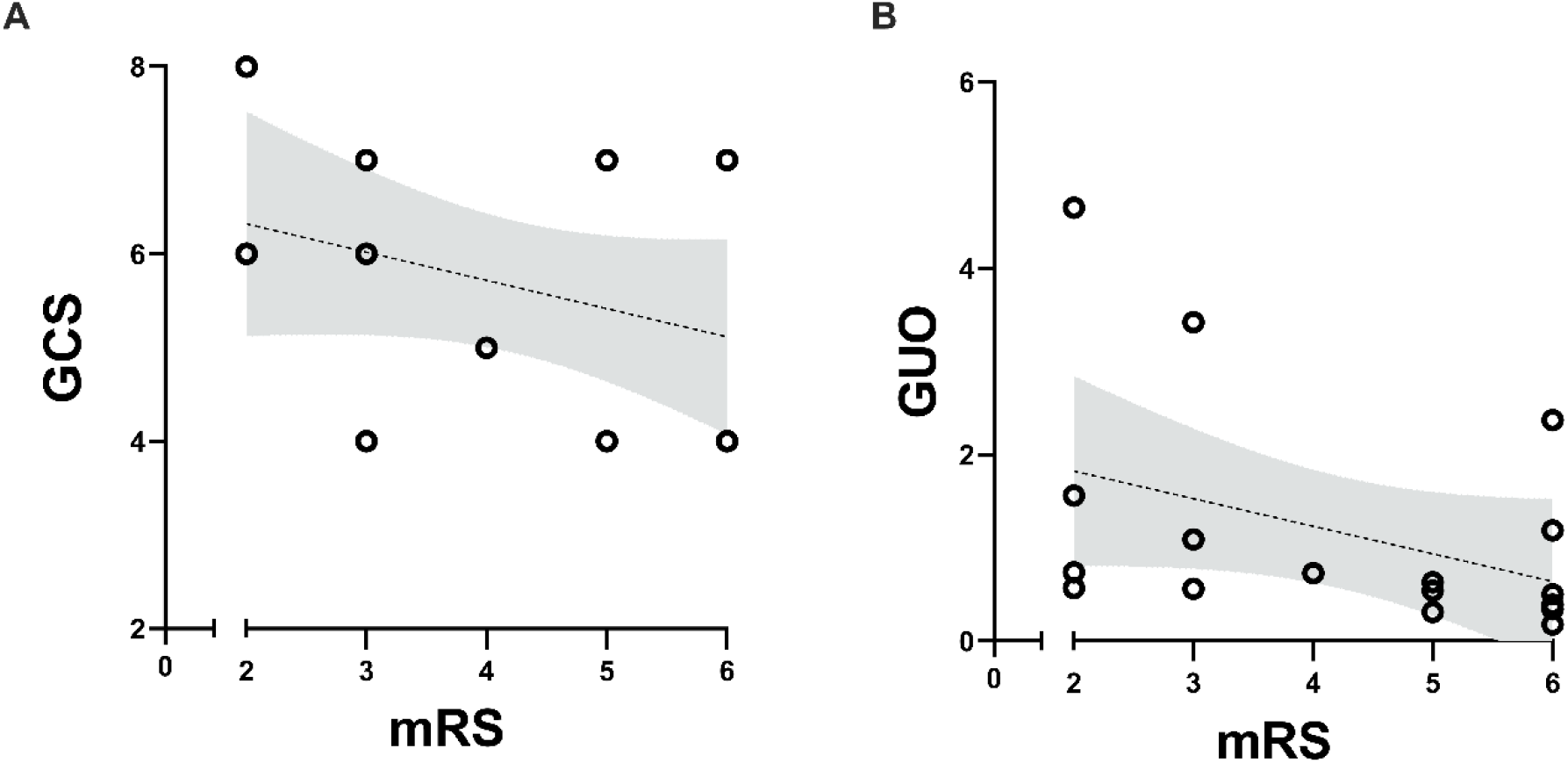
Spearman bivariate correlation between CSF purine levels and clinical neurological outcomes. **(A)** Glasgow Coma Scale (GCS) at admission did not present a statistically significant correlation with two years modified Rankin Scale (mRS) (*p* = 0.2070, *r* = −0.302, n= 17). **(B)** Guanosine (GUO) levels at admission were statistically correlated with disability 2 years after discharge (mRS) (*p* = 0.0428, *r* = −0.506, n = 17) (Survival n =11, Non-Survival n=6). Lines indicate linear regression model, and shaded area indicates the of 95% confidence intervals.

### Severe TBI disrupts CSF purinomic networks

Correlations between purines were assessed to estimate functional purinomic networks. Briefly, a stepwise metabolism of ATP generates ADP and AMP, and downstream metabolites, meaning that correlations between these molecules represent physiological interconnections. Here we show that TBI patients displayed a distinct profile of associations from controls, thereby suggesting a rupture in the physiological purinomic networks (Figure 4A and B).

**Figure 4.**
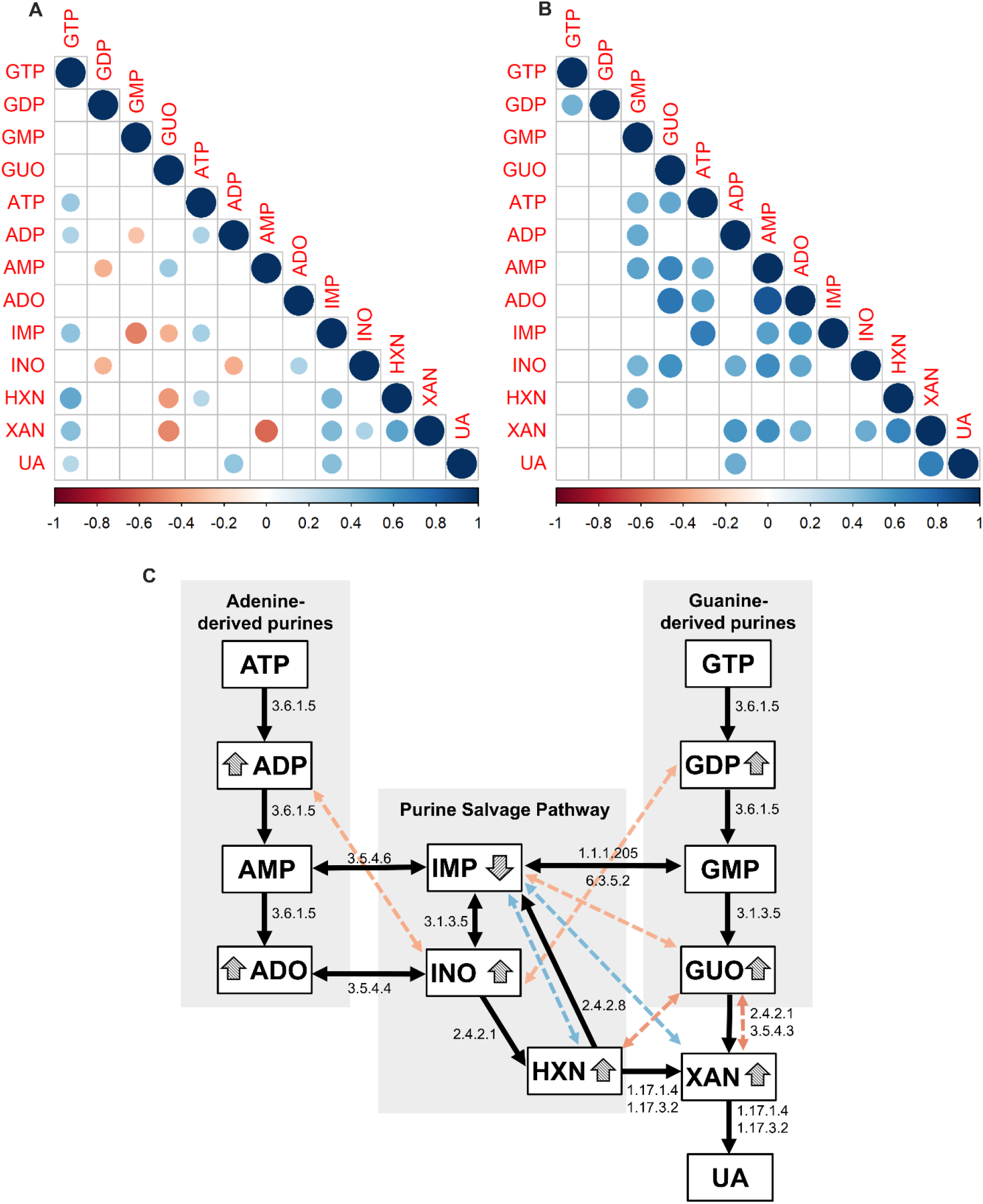
Multivariate correlations between purine derivatives in the CSF. **(A)** Spearman r correlations between purines in controls. The positive and negative correlations between adenine and guanine derivatives, and it’s products of downstream degradation suggest an integrated physiological metabolic network. **(B)** Only positive correlations between purine derivatives were observed in TBI patients, with limited association between adenine and guanine derived metabolites, and a rupture of the proposed physiological negative correlations. **(C)** Integrative model based on classical metabolic pathways of purine degradation indicated by the black flowchart, accompanied by the E.C number of the enzymes related to the respective metabolic steps leading from GTP and ATP to uric acid (UA) formation. The colored dotted double-headed arrows indicate physiological correlations disrupted by TBI. The specific purine derivatives modified by TBI are highlighted by arrowheads. **(A)** and **(B)** report Spearman’s *r* values adjusted by Bonferroni, only for values where *p* ≤ 0.05. In all figures colors indicate the nature and scale of the correlation according to the spearman *r* value, with color scales ranging from −1 (red) to +1 (blue), as indicated in **(A)** and **(B)** (Controls n= 51, TBI n = 17).

Additionally, we propose a model based on the classical purine metabolism cascade, as described in the KEGG (Kyoto Encyclopedia of Genes and Genomes) pathways. In this model we showed that bivariate correlations between purines in control subjects match the stepwise cascades of the classic purine degradation and salvage pathways. For instance, a negative correlation between ADP and GDP with INO, is observed in control subjects. This finding fits the physiological degradation of ADP and GDP driving the formation of INO. However, in TBI patients such correlations are lost, as indicated in our model by the colored dashed lines (Figure 4C).

## Discussion

This study provides the first evidence that examination of purine levels in CSF in the acute setting after TBI may serve as a promising biomarker in predicting outcome. Such “purinomics” approach showed that CSF GTP levels assessed within a short time after ICU admission appears to be a prognostic indicator of death and survival outcomes. Also, guanosine levels were associated with the impairment in a functional neurological score (mRS) two-years after ICU admission. Correlations between ATP- and GTP-derived purines, resulted in a mechanistic model that highlighted the impact of TBI on the enzymatic degradation of purines.

There is rapidly increasing interest in examining body fluids in search of biomarkers that diagnose pathological changes and predict outcomes after TBI (Atkinson et al., 2001; Zetterberg et al., 2013). The purinergic system has established components in the brain that share specific localization and functions with glutamatergic tripartite synapse, microglia, and oligodendrocytes (Burnstock, 2020, 2017). Among those components, are transporters, receptors, and Ecto-NTPDase (E-NTPDase) enzymes, which at mechanistic level control the balance of extracellular purine levels, and consequently, their effects on specific extrasynaptic and synaptic players.

We have previously shown that after a single convulsive seizure in a rat model, CSF adenine and guanine E-NTPDase activities increased in a similar time profile to classical biomarkers of neuronal (NSE) and astrocyte (S100B) death, implying that such increased activities also reflected neural cell damage (Cruz Portela et al., 2002; Oses et al., 2007, 2004). Similarly, pentylenetetrazol-kindling rats showed alterations in adenine and guanine enzymatic degradation along with increased CSF GTP, GDP, ADP, and uric acid levels (Oses et al., 2007). Notably, it has been described that TBI promotes disturbances in purinergic signaling that may favor the development of epilepsy (Boison, 2008; Englander et al., 2003; Fedele et al., 2005; Jackson et al., 2016). This last result suggests that an acute alteration in purines after TBI, could influence neurological outcomes.

The findings of the present study demonstrate that several extracellular purines are altered after a severe TBI in patients, and may serve as brain biomarkers of both short- and long-term injury caused by mechanisms associate with glutamatergic excitotoxicity (Stefani et al., 2017). Remarkably, an increased extracellular purine catabolism has the potential to generate endogenous neuroprotective anti-glutamatergic responses through adenosine and guanosine (Dunwiddie and Masino, 2001; Schmidt et al., 2000), and an antioxidant defense through the uric acid. However, considering the extravasation of active biomolecules caused by the rupture of cell membranes and axons (Johnson et al., 2013) in the first hours after TBI, it is unlikely that these endogenous neuroprotective mechanisms are ready-to-use and overcome the biochemical components of secondary damage (Stefani et al., 2017). Accordingly, exogenous administration of guanosine after mild TBI in rats displayed neuroprotective properties (Courtes et al., 2020).

While clinical studies investigating purine levels in biological fluids after TBI are scarce, previous published reports in rodent TBI (Bell et al., 1998; Marklund et al., 2006; Verrier et al., 2012) have demonstrated acute increases only in ADP, adenosine, inosine and hypoxanthine, which are consistent with the concentration profile found in our current study (Fig 1E, F, H and I respectively). However, we did not find reports of changes in GDP, guanosine, and xanthine levels in the biological fluids of patients, or in preclinical models of TBI. Hence, this work primarily demonstrates that CSF levels GDP, guanosine, ADP, adenosine, inosine, xanthine, and hypoxanthine increased above the control levels within the first 3 hours after TBI, whereas IMP was consistently decreased. Although this composite of purinergic abnormalities likely reflects damage to the brain cells, it further requires associations with clinical and neurological endpoints to be featured as a feasible biomarker in the ICU (Atkinson et al., 2001).

Among the adenine- and guanine-derived purines GTP, GDP, IMP, and xanthine were significantly increased in non-surviving compared to surviving patients. Despite IMP was decreased in the TBI group relative to control, comparison between non-surviving and surviving patients within the TBI group displayed differences (see Figures 1 G and K). The discriminatory profile of these particular purines may encompass features with potential validity to be explored as predictive biomarkers (Cristofori et al., 2005; Laketa et al., 2015).

A previous case report investigating energy failure and oxidative damage in one severe TBI patient showed a time profile for CSF adenosine, hypoxanthine, xanthine, and uric acid, that increases up to 100 h after ventricular catheter insertion. The concentrations increase 4- to 6-fold 72 h after catheter insertion for hypoxanthine, xanthine, and uric acid, and further increase before brain death, which also suggest a putative prognostic value (Cristofori et al., 2005). Albeit the time of catheter insertion and CSF collection differs from our work, we found similar increments in xanthine levels before brain death. Also, our work further highlighted the discriminatory profile of GTP, GDP, IMP, and xanthine, which expand the opportunity to identify candidate biomarkers and clinical associations.

Regarding the sensitivity and specificity of GTP, GDP, IMP, and xanthine in determining the short-term neurological prognosis of severe TBI patients, interestingly, only GTP levels displayed predictive value relative to mortality within 3 days after TBI (sensitivity of 66.67% and specificity of 100%). A potential mechanism underlying these properties has been identified in a post-mortem examination of the brains of TBI patients. The GTP binding receptors RhoA and RhoB expression levels were acutely increased after TBI, which was sustained for months after the head impact (Brabeck et al., 2004). These receptors participate in cerebral responses to injuries, regulating cerebrovascular tonus, astrocytic and microglial activation, axonal regeneration, and neuroplasticity (Stankiewicz and Linseman, 2014). In animal models of TBI, it has been reported an increase in downstream GTP-binding receptor pathways such as the RhoA-ROCK cascade, which is known to exacerbate neuronal death (Dubreuil et al., 2006; Labandeira-Garcia et al., 2017; Rikitake et al., 2005; Sabirzhanova et al., 2013). Such role is apparently corroborated by the increased GTP levels observed in non-surviving patients in our study. In addition, we sought for associations between purine levels and long-term neurologic disability. Our data demonstrates that only guanosine levels at admission in the ICU were inversely associated with mRS scores two years after TBI. Exogenous administration of guanosine has well-recognized effects against glutamatergic excitotoxicity and mitochondrial dysfunction, important components of secondary damage following TBI (Dobrachinski et al., 2019; Gerbatin et al., 2017, 2019). Based on these properties, we posit that even a build-up of endogenous levels of guanosine at admission may attenuate the progression of neuronal damage and functional decline following severe TBI.

Here, we further attempted to uncover the mechanism of purine degradation after TBI using mathematical modelling based on multiple stepwise correlations. Primarily, we showed that purine metabolism in controls is tethered to the classical enzymatic degradation steps. The correlation network present in controls reflects an integrated enzymatic degradation, endorsing the biological plausibility of these proposed associations. However, many of these correlations present in control group disappear in TBI group, suggesting an uncoupling of the enzymatic steps of purine degradation, leading to the accumulation of specific purine derivatives. For instance, in the TBI group, negative correlations (Figure 4C, red double-headed arrows) between inosine and ADP, guanosine and xanthine are lost, as well as the positive correlations (Figure 4C, blue double-headed arrows) between IMP and both, hypoxanthine and xanthine. The loss of negative correlations was accompanied by the accumulation of GDP, ADP, inosine, xanthine, and guanosine, which suggest impaired enzymatic activities, including E-NTPDases.

Noteworthy, the loss of the positive correlations between IMP and both, hypoxanthine and xanthine paralleled with a decrease in IMP levels, and consequent accumulation of hypoxanthine and xanthine. A previous study demonstrated that changes in ectonucleotidase activity in the rodent cortex, hippocampi, and caudate nucleus caused reduced AMP hydrolysis, but not ATP hydrolysis 4 hours after injury, emphasizing that purine degradation enzymatic activities are actually altered by TBI (Bjelobaba et al., 2009). Also, accumulation of xanthine and hypoxanthine in both CSF and serum, and alterations in xanthine oxidase activity have been previously reported in preclinical studies of TBI, supporting the mechanistic involvement of purine metabolism (Laketa et al., 2015; Solaroglu et al., 2005; Tayag et al., 1996). Overall, our mathematical model in CSF of patients suggests that the imbalance of purine metabolism was likely mediated by altered enzymatic activity caused by TBI. These collective findings support further investigations of key effectors mediating purine degradation after TBI, as well as the role of specific enzymes on clinical outcomes.

Our study presents limitations that should be disclosed. This is a unicentric study, that enrolled primary 20 patients with severe TBI. The low number of TBI patients, limits the power of the clinical associations with purine levels. Nonetheless, this cohort was previously explored for well-established biomarkers such as NSE, GFAP and S100B (Böhmer et al., 2011); and in spite of small sample, we replicated studies with large TBI populations. Therefore, considering the primary nature of this exploratory study, and the novelty of these findings, the results presented here reveal potentialities that needs to be better explored in further studies. Also, the comparison between TBI and controls could be biased by the location of CSF collection, that differed between controls and TBI patients. Whereas the TBI patients had their CSF samples collected from external ventricular drains, controls had lumbar puncture for CSF collection, which could affect the composition and content of various metabolites though a specific influence on purines is not known yet (Brandner et al., 2013; Hegen et al., 2018; Podkovik et al., 2020).

## Conclusion

In this work, severe TBI imprinted in the CSF of patients, particular purine-derived signatures. We identified GTP as a sensitive and specific predictive biomarker of mortality, and guanosine as an indicator of long-term functional disability.

## Data Availability

All data produced in the present study are available upon reasonable request to the authors

## Acknowledgements

The authors would like to thank the Center for Brain Injury and Repair, University of Pennsylvania (CBIR-UPENN) for the technical and scientific support, the Brazilian Agencies/Programs FAPERGS #1010267, FAPERGS/PPSUS#17/2551-0001, FAPERGS/PRONEX#16/2551-0000499-4, Programa de Internacionalização de Ciência FAPERGS/CAPES #19/25510000717-5, Program Science without Borders CNPQ #4011645/2012-6, and CNPq INNT #5465346/2014-6. Also, this research was made available with the support from National Institutes of Health grants R01NS092398, R01NS038104, R01NS094003, R01EB021293, Paul G. Allen Family Foundation, and the PA Consortium on Traumatic Brain Injury 4100077083.

## Author’s contributions

NRS, MAS, JPO, DHS and LVP built the study concept. MAS and AEB were involved in the patients’ clinical management and collected the samples and data. LVP, NRS, JPO and MAS designed the study and investigation. MSR, AK, GH, VGO, JVP and ETS prepared samples, performed the HPLC procedures, analyzed the data and created the figures. NRS and LVP wrote the first draft of the manuscript, and all authors reviewed and edited the final manuscript. DHS and LVP acquired funding.

