## Supplementary figures and images for "Cerebrospinal fluid purinomics as a biomarker approach to predict outcome after severe traumatic brain injury"

### Supplemental Figure 1

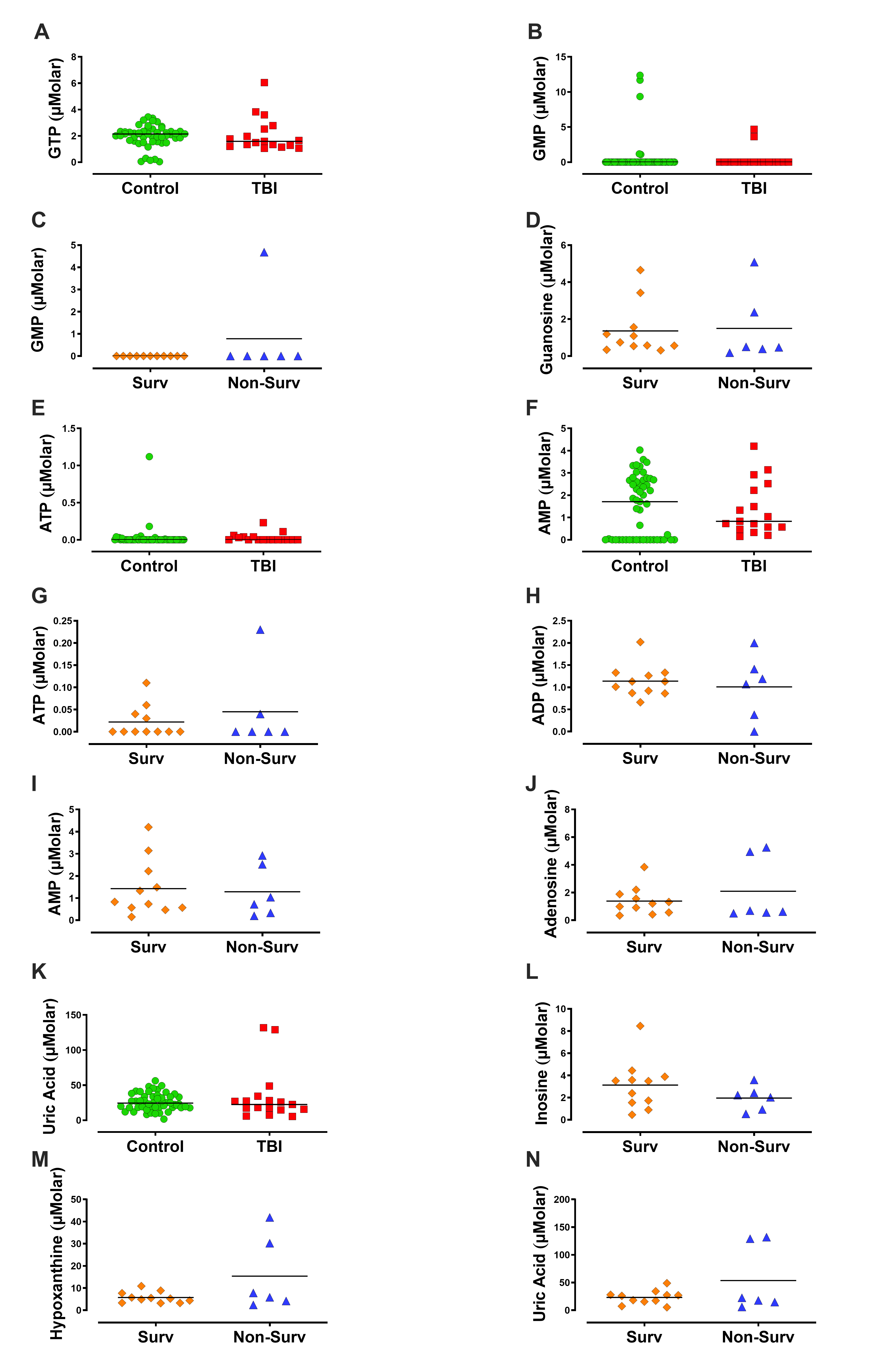
